# Fragmented criticality of infectious disease epidemics

**DOI:** 10.64898/2026.09.08.26362507

**Authors:** Boxuan Wang, Eugenio Valdano

## Abstract

Population vulnerability to an infectious disease epidemic is commonly summarized by system-level indicators such as the epidemic threshold: a single critical boundary. Yet transmission is heterogeneous across communities, host groups and transmission pathways, and public-health decisions often require identifying which parts of the system become vulnerable, and under which conditions. Here we show that epidemic criticality can itself be fragmented across population structure. Using multitype branching processes, we identify singularities governing the expected size of outbreaks that ultimately become extinct and show that coupling between population strata transforms their individual thresholds into complex-valued critical points. Their real parts locate critical changes along the transmissibility axis, their imaginary parts determine their strength and smearing, and their modes identify the subpopulations involved. In Italy, this framework reveals localized vulnerability to respiratory-pathogen emergence and can improve vaccine allocation over importation-based strategies. For measles in Texas, it identifies spatial units more homogeneous in observed outbreak burden than standard administrative or metropolitan partitions. In a One Health model of livestock-associated MRSA, it separates occupational and human–animal transmission pathways. Epidemic vulnerability is therefore organized by a structured critical landscape rather than a single threshold, and resolving this landscape can directly inform public-health risk assessment and intervention.

## Introduction

Population vulnerability to infectious disease epidemics is commonly summarized by system-level indicators such as the epidemic threshold, which identifies the transmissibility above which sustained spread becomes possible [1, 2]. The epidemic threshold has supported risk assessment and control across a broad range of epidemic systems [3–9]. This scalar description, however, compresses epidemic dynamics on heterogeneous population structures into a single critical boundary. Contemporary public-health decision-making instead requires more detailed information: which communities, host groups or transmission pathways become vulnerable first, whether vulnerability is concentrated or distributed across the population, and how these patterns change as transmissibility varies. Population structure is therefore relevant not only because it shifts the global conditions for epidemic growth, but because different parts of the system may undergo distinct changes in their capacity to sustain transmission.

Structured epidemic models already capture several aspects of this heterogeneity. Next-generation operators provide the system-level condition for epidemic growth and can quantify the contribution of particular groups or interventions to it [10–12]. Network and metapopulation approaches extend this framework to heterogeneous contacts, mobility and disease histories [3, 6, 13–16], while local reproduction metrics characterize spreading conditions associated with particular communities or introductions [17–19]. These approaches either identify a global instability or evaluate local epidemic risk at a prescribed transmission regime. They do not, however, determine whether different substructures undergo distinct critical changes as transmissibility varies. The missing object is a description of how criticality itself is distributed across a structured population.

Related phenomena occur in heterogeneous physical systems. Quenched disorder and mesoscopic structure can generate Griffiths phases and smeared transitions, while localization can cause different network regions or layers to become active at distinct parameter values [20–28]. These results show that structured systems need not be organized by a single collective transition. They have, however, been developed mainly for endemic or quasistationary spreading processes on networks with sizes approaching the thermodynamic limit. Their applicability to public-health decision-making is therefore limited, because epidemic emergence and establishment are inherently stochastic and involve small, finite numbers of infections and possibly populations. In this regime, both the location of the initial introduction and the amplification of small transmission chains become epidemiologically relevant.

Here we introduce and develop the theory of *fragmented criticality* for epidemics spreading in structured populations using multitype branching processes. We study the singularities governing the expected size of outbreaks that ultimately become extinct. In a population of isolated strata, each stratum has its own epidemic threshold. When transmission between strata is introduced, these thresholds are not simply erased into a single system-level transition: except for the global epidemic threshold, they move into the complex plane of the epidemiologically meaningful transmissibility parameter. This construction has an analogy with singularities outside the physical parameter domain that organize observable behavior, such as Yang–Lee zeros and resonance poles [29, 30]. The real part of each complex critical point identifies the transmissibility scale of the associated change, its imaginary part controls its strength and smearing, and its critical mode identifies the population strata involved. Together, these quantities define a fragmented critical landscape across population structure.

We apply this framework to three epidemiological systems. For a directly transmitted respiratory pathogen in Italy, such as influenza, SARS-CoV-2, or a newly emerging virus, fragmented criticality reveals localized regional vulnerability and can improve vaccine allocation over a strategy based on importation risk alone. For measles in Texas, critical modes identify spatial units that are more homogeneous in observed outbreak burden than standard administrative or metropolitan partitions. In a One Health model of livestock-associated methicillin-resistant *S. aureus*, they separate distinct occupational and human–animal transmission pathways. Fragmented criticality therefore provides a way to resolve when, where and through which parts of a structured population epidemic vulnerability emerges.

### Derivation of the fragmented criticality

We model epidemic dynamics in a structured population using a multitype Galton-Watson branching process [19, 31, 32]. We consider a population partitioned into *N* strata and define the stochastic variables *X*_*i→j*_ ∈ ℕ as the number of secondary infections generated in stratum *j* by an infected individual from stratum *i*. The reproduction operator **R** is defined through the expectation values of these stochastic variables: *E*[*X*_*i→j*_] = *R*_*ji*_ [33]. We parametrize it as **R** = *r***C**, where **C** is normalized so that its largest eigenvalue is equal to one, and the reference reproduction ratio *r >* 0 controls the long-term epidemic dynamics [10, 33].

We assume that *X*_*i→j*_, *X*_*i→k*_ are independent if *j ≠ k*, and that secondary infections are Poisson distributed, *X*_*i→j*_ ~ Poisson(*R*_*ji*_). This is reasonable, as transmission heterogeneity is often represented through population stratification and contact structure [2, 14]. Nonetheless, in Supplementary Methods Section 1.1 we extend the theory to arbitrary offspring distributions to account for possible individual-level overdispersion [31].

Let *p*_*i*_ be the epidemic probability, i.e., the probability that one initial infection in stratum *i* leads to a major epidemic, defined in the branching-process framework as an outbreak that never goes extinct. Under the above assumptions, the *p*_*i*_ obey the following equation [19]:

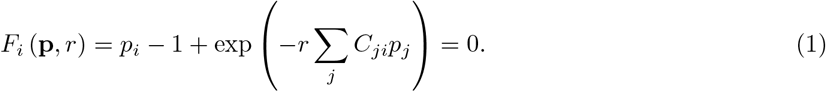

The epidemic threshold of the system is the value *r*^*c*^ of *r* such that, for any *r > r*^*c*^, *p*_*i*_ *>* 0 for at least one stratum *i*. Equivalently, it separates the regime in which extinction is certain (*r < r*^*c*^) from that in which introductions can lead to major epidemics (*r > r*^*c*^). The emergence of major outbreaks can also be characterized through the size of minor outbreaks, i.e., outbreaks that eventually go extinct. These can be studied by conditioning the branching process on eventual extinction. The conditioned process is itself a branching process with the same offspring distribution (Poisson) and tilted reproduction operator 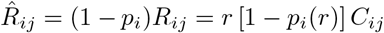 (proof in Sec. A.1). The expected size 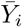 of a minor outbreak seeded in *i* obeys the equation 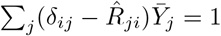 (proof in Sec. A.2), whose solution is

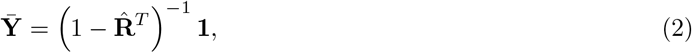

where **1** is a column vector of unit entries and 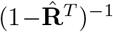 is the resolvent. We find that the epidemic threshold can be characterized as the value *r >* 0 at which the resolvent becomes singular. This occurs when 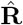 has an eigenvalue equal to one. At *r* = *r*^*c*^, the largest eigenvalue of **R** equals one by definition and **p** = 0, so that 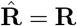 and its largest eigenvalue is also equal to one. For *r > r*^*c*^, conditioning on eventual extinction causes the largest eigenvalue of 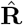 to be smaller than one. Thus, *r*^*c*^ is the unique real value at which the resolvent is singular, meaning that the epidemic threshold is the transmissibility value where the expected size of minor outbreaks becomes infinitely large. To be precise, the uniqueness requires that the inter-stratum network be strongly connected. If not, isolated subsystems may have different thresholds [33]. This interpretation of *r*^*c*^ characterizes the emergence of major outbreaks as a transition from finite minor outbreaks and, as we show below, also exposes the limitations of the epidemic threshold as a descriptor of spatially structured epidemic dynamics. For an eigenvalue *λ* of 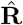 approaching one, the resolvent is 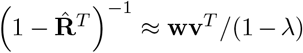, where **v, w**^*T*^ are the right and left eigenvectors of 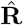, normalized such that Σ_*i*_ *v*_*i*_ = 1 and **w**^*T*^ **v** = 1. The associated critical mode is therefore localized along **w** (right eigenvector of **R**^*T*^), and the divergence is strongest in strata with large *w*_*i*_. Figure 1a–c illustrates this mechanism in a simplified two-stratum model. There, the epidemic threshold is marked by a singularity in the expected minor-outbreak size that is entirely localized in one stratum (stratum 2 in Fig. 1c). Consequently, introductions into the other stratum are extremely unlikely to lead to major outbreaks over a finite range of *r* even above the epidemic threshold (Fig. 1b,c). Critical behavior is therefore fragmented across strata, and the epidemic threshold alone is a poor predictor of introduction-specific epidemic risk. At what value of *r* do introductions into the latter stratum begin to generate major outbreaks? Fig. 1c shows that the expected size of minor outbreaks seeded in this stratum remains finite for all *r > r*^*c*^, but exhibits a pronounced peak. Numerical simulations indicate that introductions can indeed generate major outbreaks beyond this peak, but not before. This simplified model therefore identifies Eq. 2 as the key quantity for characterizing introduction-dependent critical behavior beyond the epidemic threshold.

**Fig. 1.**
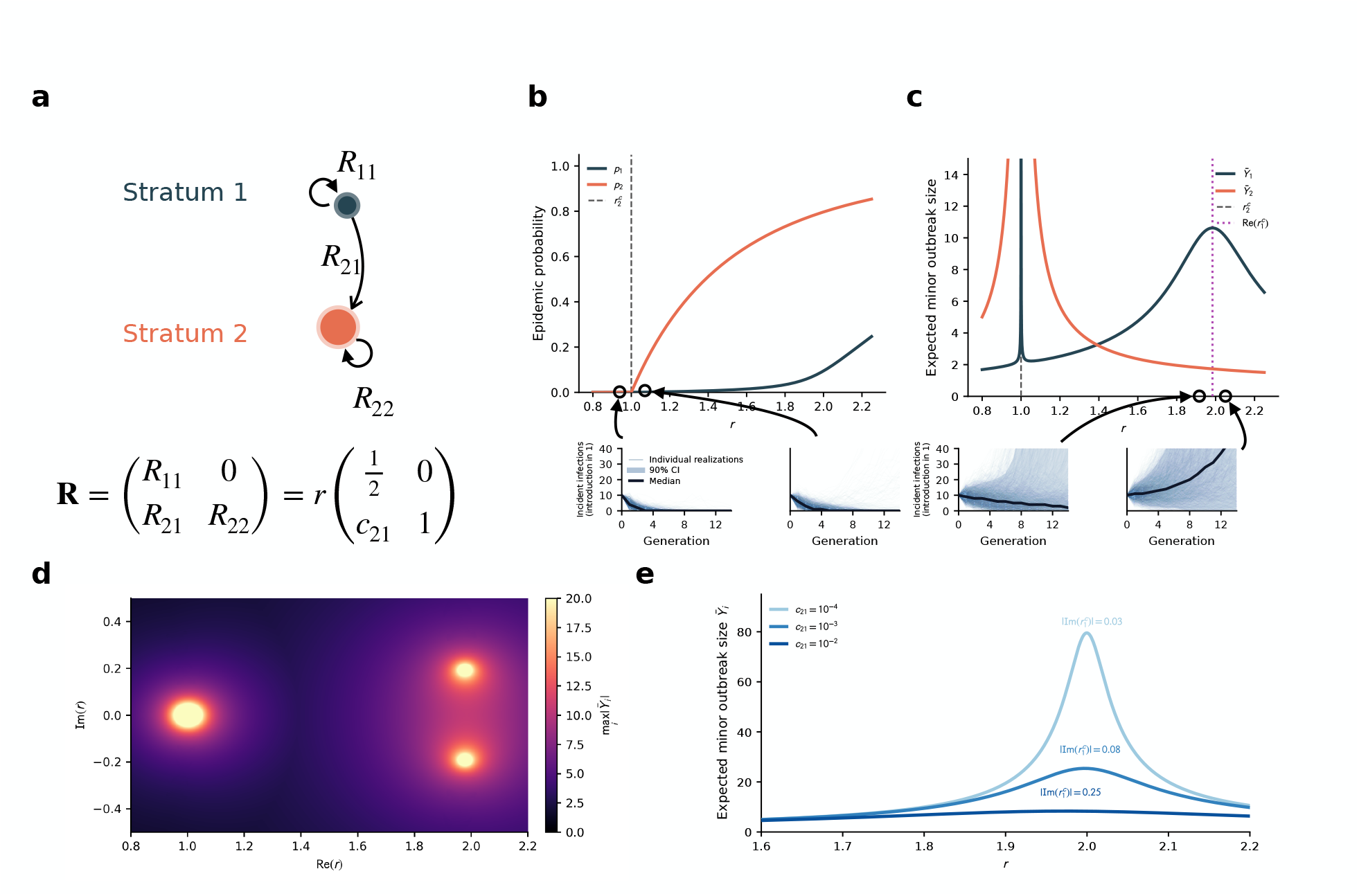
Emergence of fragmented criticality in a simplified, two-stratum model. This simplified model illustrates fragmented criticality. The two strata are labeled as 1 and 2. Both have a nonzero local reproduction ratio (*R*_11_ *>* 0, *R*_22_ *>* 0). Stratum 1 can generate infections in stratum 2: *R*_21_ *>* 0, while stratum 2 generates no infections in 1: *R*_12_ = 0. We parametrize it as **R** = *r***C** with *C*_11_ = 1*/*2, *C*_22_ = 1. *C*_21_ is set to *C*_21_ = 6 · 10^*−*3^ in panels **b, c** and **d** and explored in panel **e**. The system is weakly coupled: *C*_21_ ≪ *C*_22_. The epidemic threshold is *r*^*c*^ = 1. **a**, Schematic illustration of the model and its reproduction operator (graph representation and matrix parametrization). **b**, In the main plot, epidemic probabilities (y-axis) given introduction in stratum 1 (*p*_1_) and 2 (*p*_2_) as a function of the reference reproduction ratio *r* (x-axis). Given that the graph representation is acyclic, Eq. (1) can be solved explicitly: 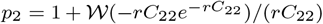 and 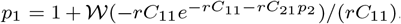. The two insets show 2, 000 stochastic epidemic realizations seeded in stratum 1 (single realizations in blue, median in black), with *r* slightly smaller and slightly larger than the epidemic threshold of the system. Quick extinction is likely in both cases. **c**, In the main plot, expected size of minor outbreaks (y-axis) given introduction in stratum 1 and 2, as a function of the reference reproduction ratio *r* (x-axis). Given that *p*_1_, *p*_2_ can be computed explicitly, also Eq. (2) can be computed explicitly in this system. The dashed black line is the epidemic threshold 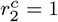 and the purple dashed line is the real part of the second critical point 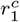 (nonreal singularity of 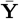, see next panel). The two insets show, as in panel **b**, 2, 000 stochastic epidemic realizations seeded in stratum 1 (single realizations in blue, median in black), with *r* slightly smaller and slightly larger than Re 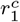. They show a change in trend, from expected exponential decay to exponential growth. **d**, continuation of the expected size of minor outbreaks 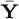 (Eq. (2)) to the complex plane of the reference reproduction ratio: *r* ∈ ℂ. Specifically, the heatmap shows the maximum between |*Y*_1_| and |*Y*_2_|, as a function of Re *r* (x-axis) and Im *r* (y-axis). It identifies one singularity in *r* = 1 ∈ ℝ (the epidemic threshold 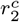) and one additional pair of complex-conjugate singularities 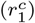. **e**, Effect of the imaginary part on the smearing and scale of the critical point. Expected size of minor outbreaks (y-axis) as a function of the reference reproduction ratio *r* (x-axis), for different values of the inter-stratum coupling *C*_21_.

To proceed, we first consider a system of isolated strata, for which **R** (and 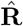) is diagonal. Each diagonal entry *R*_*ii*_ identifies a critical value of *r* 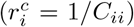 at which *R*_*ii*_ becomes equal to one and the resolvent becomes singular; the associated eigenvector is the canonical basis vector with unit entry at index *i*. Formally, the epidemic threshold of the full system is the smallest of these critical points 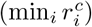. This threshold is nevertheless poorly informative because criticality is trivially fragmented: the system is simply a collection of isolated subsystems, each with its own epidemic threshold. When, instead, the off-diagonal entries *R*_*ij*_ (with *i* ≠ *j*) are comparable in magnitude to the diagonal entries, the strata are strongly coupled and the epidemic threshold captures most of the system’s critical behavior (in Supplementary Methods Section 1.2 we prove the disappearance of fragmented criticality in strongly-coupled and homogeneous populations). In the intermediate regime, however, where *R*_*ij*_ *>* 0 but inter-stratum coupling remains weak relative to intra-stratum coupling (*R*_*ij*_ ≪ *R*_*jj*_, for *i* ≠ *j*), the system should retain a “memory” of the individual stratum thresholds of the decoupled system, and indeed this memory is visible in the peak in 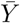 in Fig. 1c. These thresholds are no longer directly visible as singularities of the coupled resolvent, which has only one singularity for *r* ∈ ℝ_+_. Thus, although the coupled system formally has a single epidemic threshold, its critical behavior can remain fragmented, as illustrated in the two-stratum model (Fig. 1b–d). This suggests the analogy with Yang–Lee zeros in statistical physics and resonance poles in scattering theory [29, 30].

Indeed, when coupling is switched on, the *N* − 1 distinct real singularities of the decoupled resolvent (other than the epidemic threshold) move into the complex plane (Fig. 1d). Their effect remains visible on the real axis through maxima in the expected minor-outbreak size (Fig. 1c). Writing 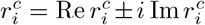, the real part locates the associated critical change along the epidemiologically meaningful transmissibility axis, whereas 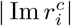 sets its smearing scale (Fig. 1e). For 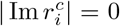 the point is a proper epidemic threshold; small 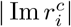 produces a high, sharp maximum, whereas increasing 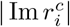 makes the response progressively lower and broader. This behavior is derived explicitly in Sec. A.4.

Because the analytically continued resolvent has conjugation symmetry, nonreal singularities occur in complex-conjugate pairs, so we consider only those with positive imaginary part. The eigenvector **w** associated with the singular mode of the resolvent quantifies the sensitivity of stratum *i* to the particular critical point. The normalized vector 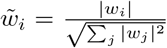 thus can be used to describe the localization of each critical point across strata. In the decoupled system, fragmentation is trivial and each critical mode is localized on a single stratum, whereas coupling can spread it across multiple strata.

The complex singularities can be identified equivalently as bifurcation points of Eq. (1). Its Jacobian is *L*_*ij*_ = ∂*F*_*i*_*/*∂*p*_*j*_ = *δ*_*ij*_ − *U*_*ij*_, with *U*_*ij*_ = (1 − *p*_*i*_)*R*_*ji*_. Since **U** and 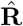 have the same spectrum, **L** becomes singular at the same points as the resolvent. Thus, the critical points correspond to values of *r* at which branches of the analytically continued epidemic probabilities *p*_*i*_ coalesce, restricting to bifurcations that are not unstable (proof in Sec. A.3). This provides an operational route to compute the fragmented critical landscape, and derive its properties, from Eq. (1).

But beyond their numerical computation, the critical points can be derived perturbatively in the weak-coupling regime (derivation in Sec. A.4). Assuming that between-stratum transmission is weaker than within-stratum transmission (*C*_*ij*_ ≪ *C*_*jj*_ for *i* ≠ *j*), as is often the case for instance in spatially structured populations [33], we obtain

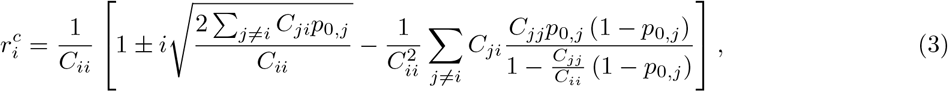

where *p*_0,*j*_ are the outbreak probability of the decoupled system, whose solutions are known: 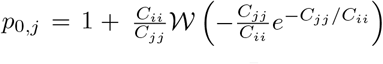, where W is the principal branch of Lambert’s *W* function. Equation (3) makes the effect of inter-strata coupling in shifting the critical point out of the real axis explicit. Its leading-order effect is to generate a nonzero imaginary part. Thus, coupling first regularizes the critical point, while its location shifts only at higher order. The localization of the critical point 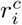 can also be computed perturbatively:

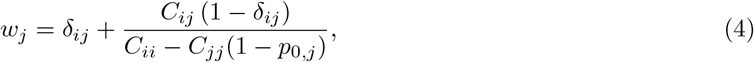

which shows that delocalization, like the shift in location, occurs at higher order than regularization. Finally, close to the critical point 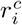, it can be shown that 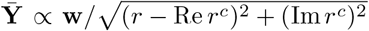, recovering the well-known resonance-like profile that underlies the analogy with scattering theory [30]. Im *r*^*c*^ regularizes the divergence, and the peak height of **Y** scales as 1*/*| Im *r*^*c*^|. The threshold is also smeared over a region with characteristic scale |*r* − Re *r*^*c*^| ~ | Im *r*^*c*^|.

The validity of the weak-coupling approximation is confirmed by the exactly solvable mean-field model presented in Supplementary Methods Section 1.3. These results also assume that, at each critical point, the critical eigenspace of the resolvent is one dimensional. This is a reasonable assumption because such degeneracies are generally unstable in data-driven systems. Nonetheless, we extend the formalism to degenerate eigenspaces in Supplementary Methods Section 1.4.

### Vaccine allocation for an emerging respiratory pathogen in Italy

We reconstructed the province-level reproduction operator of a directly transmitted respiratory pathogen in Italy from mobility-derived colocation data [33] (see Sec. A.6). The resulting critical landscape was strongly fragmented (Fig. 2a), with most modes localized on individual provinces or small groups of neighboring provinces. For example, one mode was almost entirely localized in Naples, a major Italian city (99% of 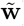), whereas another was shared between Macerata and Fermo (88% and 12%, respectively), two neighboring and relatively small provinces in central Italy. Thus, even in a connected population, the epidemiologically relevant units associated with criticality need not extend across the whole system or coincide with administrative divisions [34, 35].

**Fig. 2.**
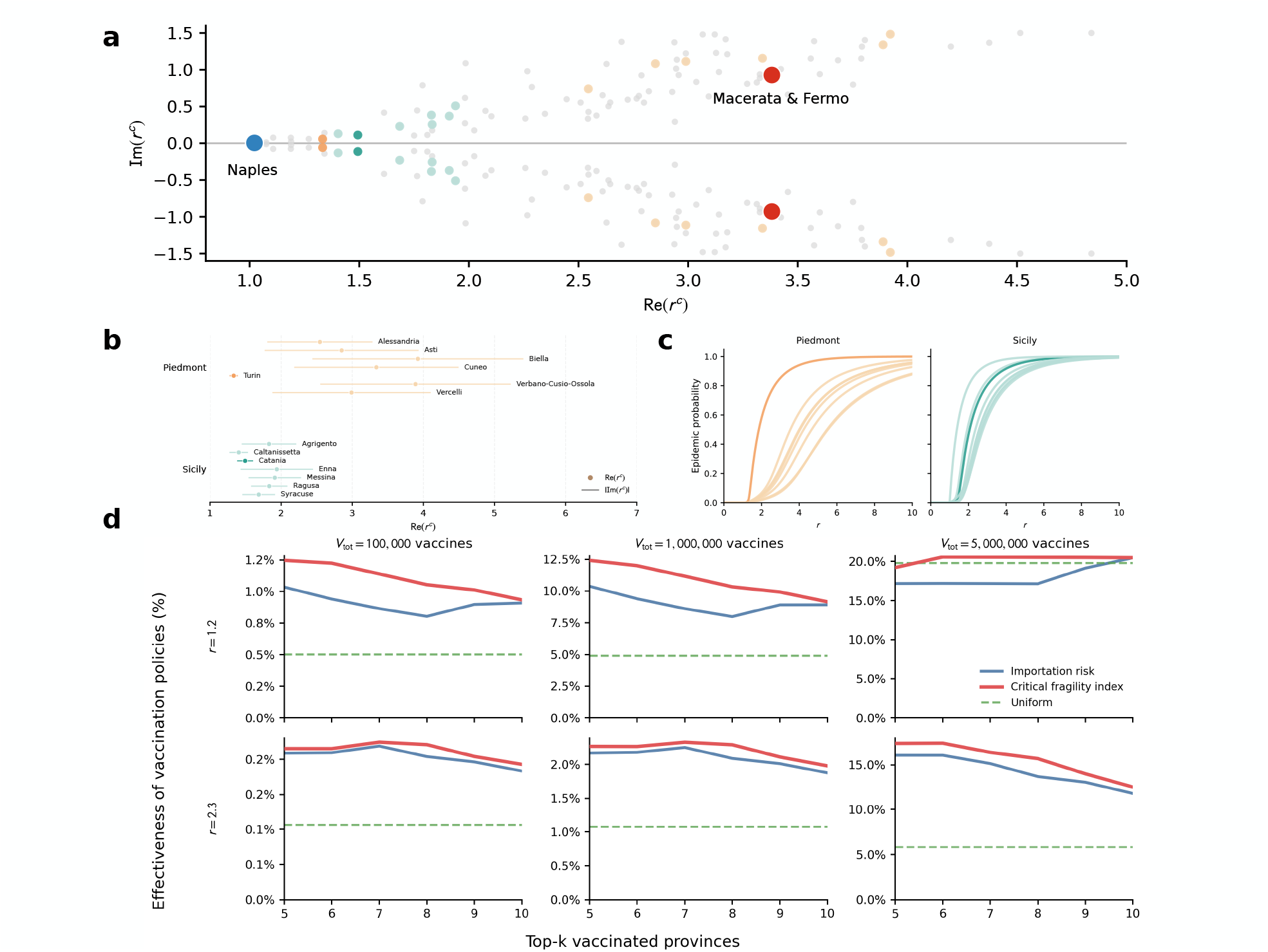
Critical points and vaccine allocation for an emerging respiratory pathogen in Italy. **a**, Critical points associated with Italian provinces in the complex *r*^*c*^ plane. Each gray point corresponds to one province-associated critical point (association established through **w**); complex-conjugate pairs are symmetric about the real axis. Coloured points indicate provinces in Piedmont (orange) and Sicily (teal). Large blue and red markers identify the critical points associated with Naples and Fermo–Macerata, discussed in the main text. **b**, Critical points associated with the provinces of Piedmont and Sicily. Dots and x-axis indicate real parts (Re *r*^*c*^) and horizontal lines indicate imaginary parts (±| Im *r*^*c*^|). **c**, Epidemic probability after introduction in individual provinces of Piedmont (left) and Sicily (right), as a function of *r*. **d**, Effectiveness of vaccination policies in the simulated experiment of preventive vaccination: uniform allocation (green dashed), allocation based on risk of importation (blue), allocation based on the critical fragility index (red). Columns correspond to vaccine stocks of *V*_*tot*_ = 10^5^, 10^6^, 5 × 10^6^ doses. Rows correspond to assumed reference reproduction ratios of *r* = 1.2 and *r* = 2.3. The horizontal axis gives the number of provinces prioritized for vaccination. The vertical axis reports the estimated effectiveness. See Sec. A.6 for methodological details.

Fragmentation also differed across regions. In Piedmont, the mode localized in its major city (Turin) had both smaller Re 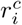 and Im 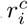 than those of the surrounding provinces, corresponding to an earlier and sharper critical change, and a centralized regional pattern (Fig. 2b). Sicily instead displayed several comparable critical modes with small imaginary parts, producing a more distributed pattern with multiple relevant critical modes. Epidemic probabilities reflected this difference: Turin separated clearly from the other Piedmont provinces, whereas several Sicilian provinces underwent comparable transitions (Fig. 2c). These structures are not captured by a single national, or even regional, epidemic threshold.

We then tested whether the fragmented critical landscape could inform vaccine allocation in a stylized invasion scenario with importations from Mexico, the origin of the 2009 H1N1 pandemic. Other importation sources gave similar results – see Supplementary Figs. S2 and S3. For a limited stock of perfectly effective preventive vaccine, we compared uniform allocation, allocation based on importation risk, and a critical-fragility ranking combining importation risk with fragmented criticality (see Sec. A.6 for methods and data). At *r* = 1.2, critical-fragility allocation substantially outperformed importation-based allocation across the vaccine stocks considered, whereas at *r* = 2.3 the two targeted strategies produced identical results because they selected the same top-*k* provinces (Fig. 2d). This simplified experiment is not an operational vaccination prescription, but shows that fragmented criticality contains intervention-relevant information beyond importation risk alone.

### Spatial scales of measles outbreaks in Texas

Fragmented criticality does not remain localized at the scale of administrative units. We explored measles outbreaks in Texas, USA, by constructing a county-level reproduction operator using Colocation Maps and vaccination coverage [36] (see Sec. A.7). We assigned each county to the critical mode to which it contributes most strongly. The resulting partition grouped counties together but is spatially coherent: counties belonging to the same mode form extended, mostly contiguous groups rather than dispersed sets (Fig. 3a). We quantified this correspondence by comparing the critical-mode partition with individual counties, larger administrative regions and metropolitan statistical areas. The agreement is highest with metropolitan statistical areas, with an F1-score of 0.60 (see Sec. A.7), compared with 0.45 for individual counties and 0.42 for administrative regions (Fig. 3b). This scale is also epidemiologically meaningful for preparedness, as metropolitan areas have been used as operational units for the rapid distribution of medical countermeasures in the United States [37–39]. Our analysis, however, revealed that they are not optimal, as we compared our theoretically derived partition with the observed geography of the 2025 Texas measles outbreak [40]. Counties belonging to the same critical mode had a higher within-group agreement in observed measles burden (0.70) than counties grouped by metropolitan statistical areas (0.62) or larger administrative regions (0.60) (Fig. 3b). The critical modes therefore identify spatial units that are not imposed a priori, yet are simultaneously consistent with functional metropolitan geography yet even more aligned to the observed epidemic burden. Fragmented criticality can thus reveal the spatial scale at which epidemic emergence is effectively organized, rather than requiring this scale to be specified beforehand. This is consistent with growing evidence that administrative boundaries can obscure epidemiologically relevant spatial structure, and that transmission and control may be better organized around functional regions emerging from mobility and local epidemic dynamics [41, 42].

**Fig. 3.**
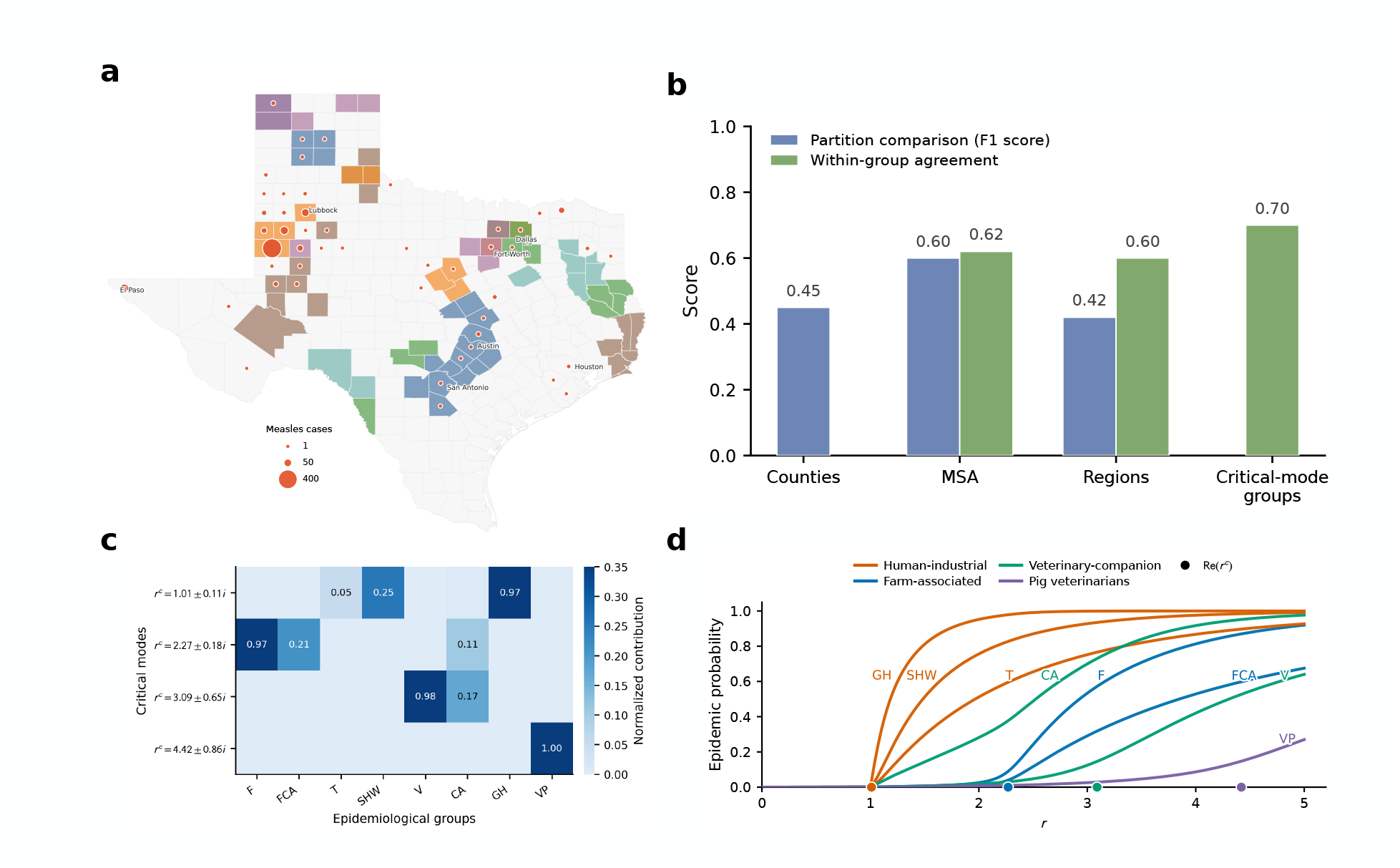
Fragmented criticality identifies spatial scales of measles outbreaks in Texas and transmission pathways of MRSA in a One Health system. **a**, Partition of Texas counties into critical modes calculated from the reproduction operator informed by spatial co-location and vaccination data from 2025. County colors identify the critical mode to which each county contributes most strongly; circle size indicates the number of reported measles cases in 2025. **b**, Comparison of the critical-mode partition with alternative spatial partitions. It reports F1-scores of the comparison with counties, regions and metropolitan statistical areas (blue), and within-group agreement in observed measles burden in different partitions: regions, metropolitan statistical areas and critical-mode partition (green). **c**, Composition of the four dominant critical modes of the LA-MRSA One Health transmission network. Columns correspond to epidemiological groups (population strata) and cell values report the normalized mode contributions. The following abbreviations are used for the epidemiological groups: farmers (F), farm companion animals (FCA), transporters (T), slaughterhouse workers (SHW), veterinarians (V), companion animals (CA), general human population (GH), pig veterinarians (VP). The modes identify four distinct transmission pathways: human-industrial (containing T, SHW, GH), farm-associated (F, FCA), veterinary-companion (V,CA), and pig-veterinarian (VP). **d**, Epidemic probability after introduction in each epidemiological group in the LA-MRSA network. Epidemiological group abbreviations and transmission pathways are the same as in **c**. Different colors correspond to different transmission pathways. Dots indicate the real part of the critical points identified in **c**.

### Transmission pathways of livestock-associated MRSA

In the previous two applications populations were structured spatially. The theory, however, applies to any kind of stratification. Here, we considered the spread of livestock-associated methicillin-resistant Staphylococcus aureus strain ST398 (LA-MRSA) in a One Health setting. We reconstructed the reproduction operator describing transmission among human, animal and occupational groups using data from Ref. [43]: see Sec. A.8 for the description of the methodology. The system included farmers, farm companion animals, transporters, slaughterhouse workers, veterinarians, companion animals, the general human population and pig veterinarians. Fragmented criticality separated this system into four distinct functional modes (Fig. 3c). The first linked transporters, slaughterhouse workers and the general human population, defining an industrial human exposure pathway. The second coupled farmers and farm companion animals; the third linked veterinarians and companion animals; and the fourth was dominated by pig veterinarians. The critical structure therefore partitioned the transmission system according to epidemiological function rather than geography. An analysis of epidemic probabilities validated this organization (Fig. 3d). The human-industrial groups became critical around *r* ≃ 1, followed by the farm-associated groups around *r* ≃ 2, the veterinarian–companion-animal groups around *r* ≃ 3, and the pig-veterinarian pathway at still larger values of *r*. Thus, the critical modes predicted both which epidemiological groups behave collectively and the order in which these transmission pathways become relevant as transmissibility increases. Fragmented criticality is therefore a property of structured transmission itself: its relevant units can be geographical communities, metropolitan systems or functional interfaces between host populations.

## Discussion

Fragmented criticality changes how heterogeneity enters the description of epidemic establishment. We show that population stratification can reorganize criticality into multiple modes, rather than simply affecting a single system-level critical indicator as in previous approaches based on the epidemic threshold. Each mode is characterized by a transmissibility scale, a degree of strength and smearing, and a localization across the structured population.

Critical modes also identify the epidemiological scale at which emergence and circulation are organized. They can remain localized on individual communities or small groups as in the case of the respiratory pathogen in Italy, aggregate into larger spatial units as in the case of measles in Texas, or identify delocalized, non-geographical transmission pathways as in the case of MRSA in the One Health application. These relevant units are therefore not fixed by administrative geography or by an imposed higher-level partition, but emerge from the critical structure of the dynamic process.

This distinction has consequences for epidemic preparedness and control. A global epidemic threshold establishes whether sustained transmission is possible somewhere in the system, but not which parts of the population are closest to a critical change or whether vulnerability is concentrated or distributed. Fragmented criticality provides this information through the location, strength and support of the critical points. The Italy vaccination experiment illustrates its potential relevance: even under a deliberately simplified intervention model, allocation informed by the critical landscape was more effective than allocation based on importation risk alone in most scenarios.

Our study has limitations. Our framework is designed for epidemic emergence, and circulation over time spans that allow modeling as a multitype branching process, i.e., over periods where the reproduction operator can be assumed to be approximately constant [33]. It does not include nonlinear effects that follow long-term epidemic circulation. Our theory is agnostic to how the reproduction operator is reconstructed, but the analyses presented here assume that changes in transmissibility scale transmission channels proportionally. This excludes changes in the contact networks that depend on circulation itself, such as behavioral adaptations. The quantitative parametrization of the reproduction operator is also only as reliable as the data used: uncertainty or bias in mobility, contact, susceptibility or vaccination data propagates to the inferred critical points and modes. Finally, the stratification used in the data determines the finest structure that can be resolved. Critical modes can combine input strata into larger functional units, as in Texas, but cannot recover heterogeneity below the resolution represented in the data used to parametrize the reproduction operator.

Fragmented criticality shifts the question posed near epidemic emergence. It is not only whether a population has crossed a global epidemic threshold, but which subpopulations, communities or transmission pathways become capable of sustaining spread, at which transmissibility values, how sharply these transitions occur, and how localized they remain. Resolving this structure provides a detailed picture of epidemic vulnerability that can inform policy. Epidemic emergence in heterogeneous populations is thus not exhausted by a scalar critical boundary: it is organized by a critical landscape whose geometry identifies when, where and how large-scale spread becomes possible.

## Supporting information

Threshold_SI

## Acknowledgments

Colocation data were available thanks to *AI For Good at Meta*. This study was partially supported by the Horizon Europe grant SIESTA (101131957) and the Agence Nationale de la Recherche (ANR) JCJC grant DiscoReel (ANR-25-CE45-5346-01).

## Author contributions

E.V. conceived the study. B.W. and E.V. designed the study. E.V. developed the theory. B.W. developed the numerical implementation and code. B.W. analyzed the data and performed the numerical analyses.

B.W. and E.V. interpreted the results. B.W. and E.V. wrote the article.

## A Methods

### A.1 Offspring distribution conditioned on extinction

Here we derive the branching process conditioned on eventual extinction. Let **X**_*i*_ be the vector with *X*_*i→j*_ as its *j*th entry. We call *f*(·|*µ*) the probability mass function of the Poisson-distributed offspring distribution, with expectation value *µ*. We call *g* its associated probability-generating function. We call 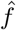, 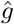 the probability mass function and probability-generating function of the extinction-conditioned process. More details in Supplementary Methods Section 1.1.

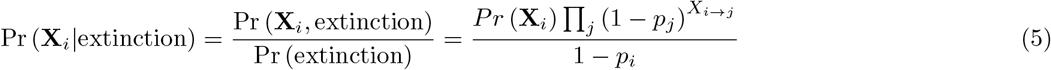

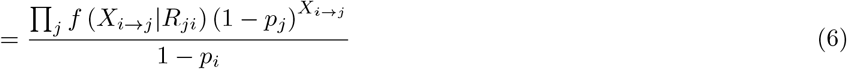

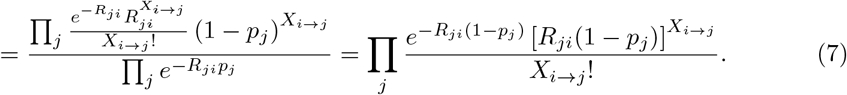

This implies that (*X*_*i→j*_|extinction) is still Poisson-distributed, with expectation value 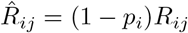.

### A.2 Expected size of minor outbreaks

Let us consider the branching process conditioned on eventual extinction. Let *Y*_*i*_(*t*) be the stochastic variable encoding the cumulated outbreak size at generation *t*, given origin (initial case) in *i*. Let *Y*_*i*_ = lim_*t→∞*_ *Y*_*i*_(*t*) be the final outbreak size. Let 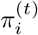 be the probability mass function of *Y*_*i*_(*t*) and 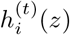 (*z*) its probability-generating function. We denote the asymptotic fixed point (*t* → ∞) values as *π*_*i*_ and *h*_*i*_, and derive a functional equation for *h*_*i*_. At *t* = 0 (initial seeding) and assuming that seeding occurs in *i*, one has 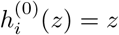. Then we consider independent, identically distributed copies of *Y*_*i*_(*t*) which we identify with the index *a*: 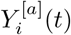: they can be seen as describing independent outbreaks. In analogy to what was done in Ref. [44] for the case of a single homogeneous population, we note that

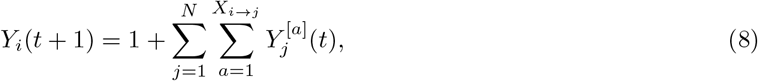

This amounts to considering each secondary infection of the initial seeding case as generating an independent outbreak, which is true according to the properties of the branching process. Note that *X*_*i→j*_, the number of infections generated by the index case in community *j*, is sampled from the offspring distribution of the branching process conditioned on extinction. Eq. (8) then states that the total outbreak size at generation *t* + 1 is the sum of the initial infected host (1) and the size of each outbreak generated by its secondary infections, measured after *t* generations.

By the properties of probability generating functions, Eq. (8) implies the following recursion relation for 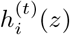:

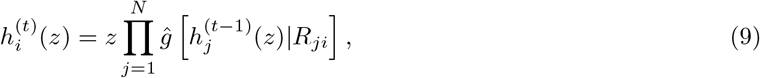

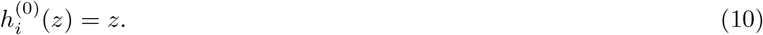

Performing the *t* → ∞ limit we get a functional equation for *h*_*i*_(*z*):

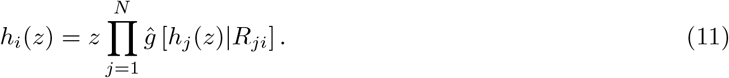

We now can use Eq. (11) to compute the expected final outbreak size 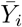, conditioned on the origin being in *i* and on extinction. To do this, we note that 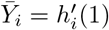 and so we derive Eq. (11) by *z* and then evaluate it in *z* = 1, remembering that 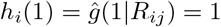. This gives the following:

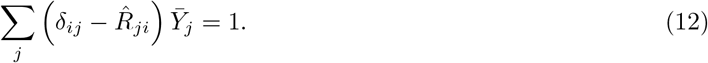

Inverting 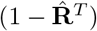, one gets Eq. (2).

### A.3 Linking bifurcation points to critical points

#### Relationship between 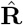 and U

Let us define **S** as a diagonal matrix with 1 − *p*_*i*_ in its diagonal entries. Then 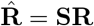 and **U** = **SR**^*T*^. **S** is clearly invertible because its diagonal entries are asymptotically zero only when *r* → ∞. Then 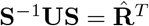, so **U**, 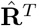 are similar and thus have the same spectrum. This implies that also **U**, 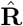 have the same spectrum.

Let **v, w**^*T*^ be the right, left eigenvectors of 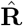 for a specific eigenvalue (assumed nondegenerate). Then, using the similarity condition, for the same eigenvalue, the right and left eigenvectors of **U** are, respectively, 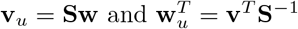.

#### Stability of bifurcation points

Equation (1) is the fixed-point equation of the dynamics describing the evolution of *p*_*i*_(*t*), the probability that an outbreak seeded in stratum *i* is still active by *t*:

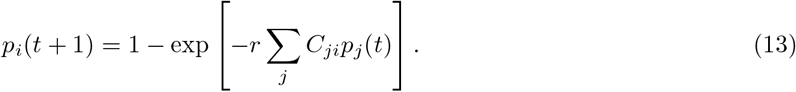

Then, *p*_*i*_ is the asymptotic probability at time *t* → ∞ (see Ref. [19]). With Eq. (13), we can study the stability of the fixed point. Writing *p*_*i*_(*t*) = *p*_*i*_ + *a*_*i*_(*t*) and expanding up to linear terms in **a**, one gets

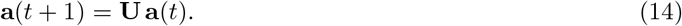

Thus, the bifurcation point **p** of Eq. (1), fixed point of Eq. (13), is stable if *ρ*[**U**] *<* 1 and unstable if *ρ*[**U**] *>* 1, where *ρ* is the spectral radius (the largest absolute value of the eigenvalues).

These results allow us to filter the bifurcation points corresponding to our critical points. The condition for having a bifurcation point (det **L** = 0) requires that one eigenvalue of 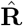 (and so **U**) be equal to one, so the case of *ρ*[**U**] *<* 1 is excluded: neither singularities of the resolvent 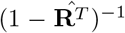 nor bifurcation points of Eq. (1) can exist. The case *ρ*[**U**] *>* 1 is also excluded by the fact that a solution of Eq. (1) which is an unstable fixed point of Eq. (13) is not an asymptotic outbreak probability. So the remaining case is *ρ*[**U**] = 1, which means that the mode becoming singular is also the one setting the spectral radius of **U**. Determining the stability of the bifurcation points at *ρ*[**U**] = 1 would require expanding Eq. (13) up to (at least) quadratic order in **a**. However, we can already assume that they have at least one basin of attraction because Eq. (13) must have at least one asymptotic solution determining the probability that an outbreak goes extinct or not, which must thus be reached by the evolution of **p**(*t*).

This completes the proof that the critical points of the epidemic dynamics, defined as the singularities of the resolvent of the expected outbreak size, are in one-to-one correspondence with the not-unstable bifurcation points of the equation describing the major outbreak probability (Eq. (1)).

### A.4 Perturbative weak-coupling solution

We consider

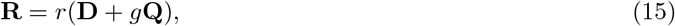

with **D** diagonal and **Q** with zero diagonal entries. Also, *ρ*[**D**] = 1 so that *ρ*[**Q**] = 1 at O(*g*). Equation (1) and its Jacobian **L** become

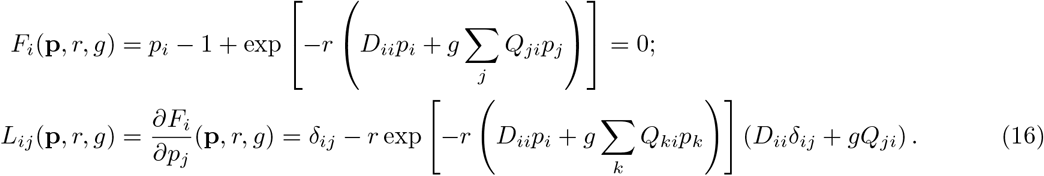

We start from the epidemic threshold of the ℓth stratum in the decoupled regime (*g* = 0). Its value is 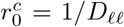 and, there, each entry of the outbreak probability vector obeys *p*_0,*i*_ = 1 − exp(−*D*_*ii*_*p*_0,*i*_*/D*_ℓℓ_). This is explicitly solvable by means of the principal branch of Lambert’s W function:

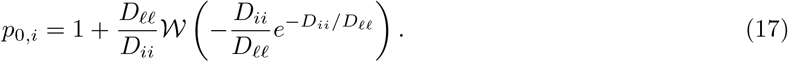

Note that *p*_0,*i*_ = 0 wherever *D*_*ii*_ ≤ *D*_ℓℓ_. The critical point in the decoupled regime is thus explicitly computable: 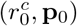. Our goal is now to find how this critical point changes when *g* is no longer zero. We assume *g* small (weak coupling) and compute the perturbative corrections to the critical point up to linear order in *g*.

The first step is to find the solution **p** = **p**(*r, g*) of Eq. (1) in the neighborhood of 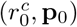. This function is not unique because, being a critical point in the decoupled system, **L** is singular there: 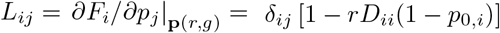 (the singularity comes from *L*_ℓℓ_ = 0). We thus need to first find the solutions in the restriction where **L** is invertible and then find the different folds of the solutions, using the Lyapunov-Schmidt reduction [45].

Given the particular structure of **L** both its kernel and its cokernel are spanned by **e**_ℓ_, vector of the canonical basis with one in ℓ and zero elsewhere. First, we decompose **p** = **p**_0_ + *ξ***e**_ℓ_ + **q** into its component in ker **L** (*ξ***e**_ℓ_) and outside of it (**q**).

Then, we focus on the part of Eq. (1) projected onto the image of **L**: Here, it simply means considering *i*≠ ℓ. We use this to find **q** = **q**(*ϵ, g, ξ*) as a function of *ϵ, g, ξ* (for convenience, we define a shifted transmissibility as 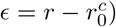).

We work up to quadratic order *ϵ, g, ξ*: **q** = *ϵ***q**_*ϵ*_ + *g***q**_*g*_ + *ξ***q**_*ξ*_ + 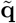, where 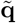 contains all quadratic powers. We use Eq. (1), restricted to *i* ≠ ℓ, to find **q**_*ϵ*_, **q**_*g*_, **q**_*ξ*_. We do not explicitly compute the term 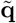 because it will play no role in the following. Combining Eq. (1) and Eq. (15), and working order by order up to linear orders in *ϵ, g, ξ*, one gets, for *i* ≠ ℓ,

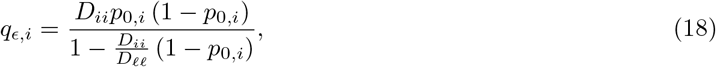

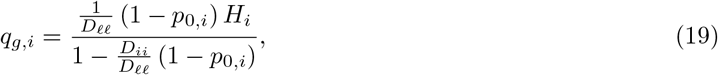

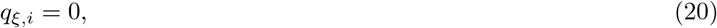

where we defined *H*_*i*_ = Σ_*j*_ *Q*_*ji*_*p*_0,*j*_.

We then move to the part of Eq. (1) projected onto the cokernel of **L**. Here, it simply means considering *i* = ℓ in Eq. (1):

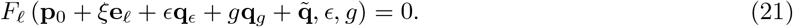

We expand up to quadratic order in *ϵ, g, ξ*. Any term arising solely from a linear expansion of the first argument of *F*_ℓ_ is zero because proportional to *L*_ℓ*j*_ = 0. This means that 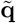 provides no contributions to the equation and that the term linear in *ξ* is also zero. The linear term in *ϵ* gives Σ_*j*_ *L*_ℓ*j*_*q*_*ϵ*,*j*_ +∂_*ϵ*_*F*_ℓ_(**p**_0_, 0, 0). The first term is zero because again proportional to *L*_ℓ*j*_. The second term is also zero because, from Eq. (16), one gets 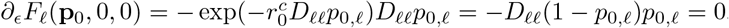 because *p*_0,ℓ_ = 0.

Then, up to quadratic order, Eq. (21) is

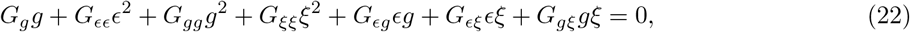

where we have introduced the relevant coefficients. Solutions of this equation in terms of *ξ* = *ξ*(*ϵ, g*) are thus the solutions **p** = **p**(*r, g*) we were looking for. Being Eq. (22) quadratic in *ξ*, there are two distinct solutions of Eq. (16) around 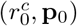, attesting to the singularity of **L**. The critical point is the bifurcation point where those equations coincide, which we find by setting the discriminant of Eq. (22) to zero and solving for 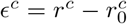 as a function of *g*. This gives

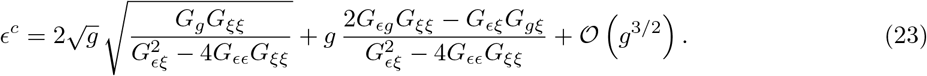

The term *G*_*gg*_ does not appear because it is associated to higher order terms. The remaining factors to be computed are thus

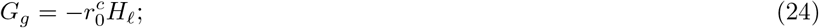

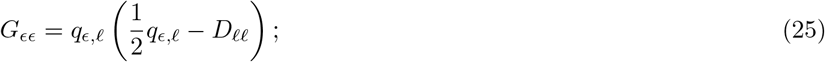

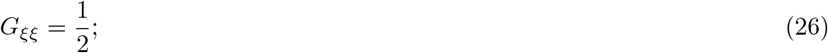

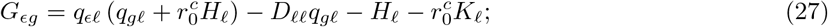

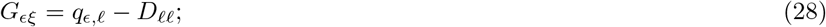

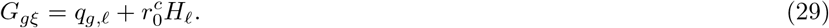

where all the derivatives are computed in the base point (*r*^*c*^, **p**_0_) and **K** = **Q**^*T*^ **q**_*ϵ*_. Using this, one gets

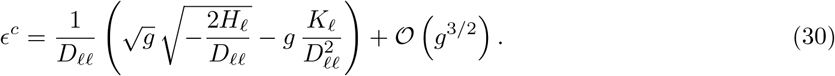

Since, clearly, *D*_ℓℓ_, *H*_ℓ_ *>* 0, the negative factor under the square roots in the 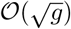 moves the critical point out of the real axis, while the *O*(*g*) term decreases its value on the “physical” transmissibility axis:

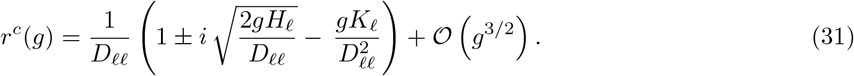

Notably, this can be written as a function of these two terms containing the between-strata coupling:

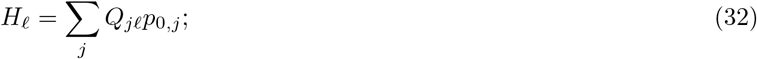

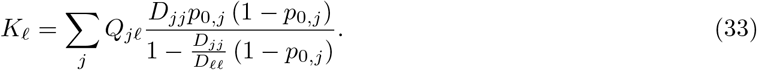

They are explicitly computable in the decoupled system, using Eq. (17). Note that in the above sums over *j* only the strata for which *D*_*jj*_ *> D*_ℓℓ_ contribute with nonzero terms, meaning that, at this order, only the strata which are above threshold in the decoupled system (at *r* = 1*/D*_ℓℓ_) contribute to shifting the threshold of stratum ℓ.

Also, note that, in weak coupling, the lowest meaningful order is 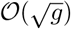, so the effect of coupling is first to smear the threshold by moving it to the complex plane, then only for higher coupling to actually displace it.

#### Localization

To study the localization of the critical point in Eq. (31) to matching order in *g*, we need to study the impact of coupling on the ℓth right eigenvector of 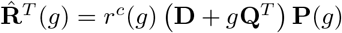, where **P** = diag(1 − *p*_*i*_), a diagonal matrix with 1−*p*_*i*_ in its *i*th diagonal entry. We need to study the eigenvector equation 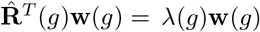. Since *r*(*g*) does not change the eigenvectors, we will study the equivalent equation

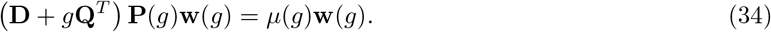

In the decoupled system, **w**(0) = **e**_ℓ_ (canonical basis vector) and *µ*(0) = *D*_ℓℓ_. We expand perturbatively: 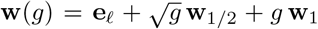 with *w*_ℓ_ = 1, *w*_1*/*2,ℓ_ = *w*_1,ℓ_ = 0, and 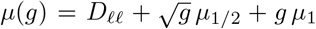. We also expand 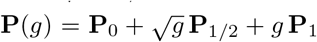. Plugging these into Eq. (34) and expanding order by order in *g*, we get the following 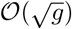 order:

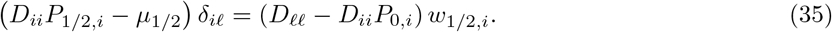

This implies 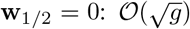 does not contribute to mixing the eigenvectors. This is not surprising given that contributions to 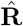 at this order are diagonal.

Order O(*g*) instead gives

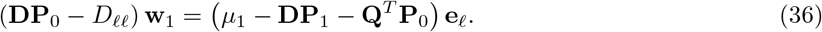

This, evaluated for *i* ≠ ℓ, gives the final perturbative expression of the eigenvector:

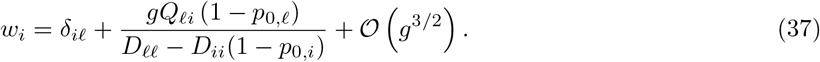

This proves that i) the delocalization of the critical point appears only at linear order in the coupling constant; the leading 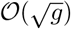 order smears it but does not delocalize it. ii) Strata contributing to the critical points are those transmitting to ℓ (*Q*_ℓ*i*_ *>* 0).

#### Interpretation of the real and imaginary parts of the critical points

We can synthetically write the equation for *F*_ℓ_ (bifurcation equation), expanded up to quadratic order (Eq. (22)), as *α*(*ϵ, g*)*ξ*^2^ + *β*(*ϵ, g*)*ξ* + *γ*(*ϵ, g*) = 0. Its solutions (the two folds) are thus 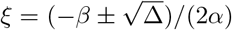. The discriminant Δ is what vanishes at the critical point *r*^*c*^ (Eq. (23)) and its complex conjugate *r*^*c\**^, so it can be written as Δ = *κ*(*g*)(*r* − *r*^*c*^)(*r* − *r*^*c\**^). Finally, deriving *F*_ℓ_ by *ξ* one gets

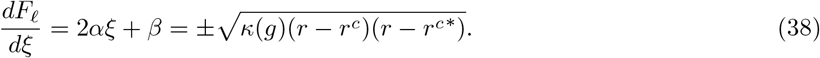

Let us call *F*_*⊥*_ the part of Eq. (16) projected outside the kernel of **L**, i.e., *i*≠ ℓ. Then

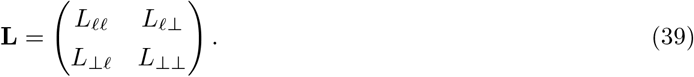

*L*_*⊥⊥*_ is invertible, so

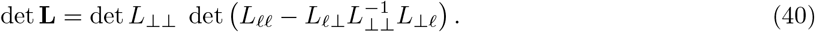

Remembering that **p** = **p**_0_ + *ξ***e**_ℓ_ + **q** and deriving Eq. (16) by *ξ* evaluated in the base point, one gets

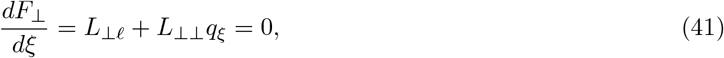

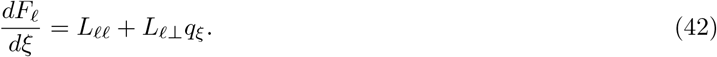

Combining Eq. (38), Eq. (40), Eq. (41) and Eq. (42) one gets that

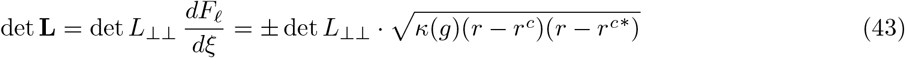

Given that the only term becoming zero in the critical point is the square root (det *L*_*⊥⊥*_ ≠ 0), the square root also determines the leading behavior of the eigenvalue of **L** becoming singular in *r*^*c*^. This means that, close to the critical point, the resolvent has the following leading-order behaviour

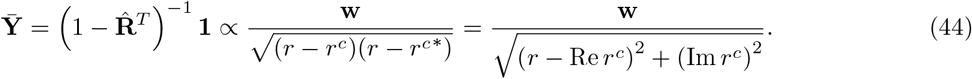

### A.5 Numerical evaluation of the critical points

To numerically compute the critical points, one needs to evaluate 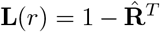, i.e., the inverse resolvent, for *r* ∈ ℂ. For a given value of *r*, first we found **p** by solving Eq. (1). We worked on a vector of dimensionality 2*N*, to account for Re **p** and Im **p**. The solution was reached first by fixed point iteration of Eq. (13) and then refined by a Newton-type solver. Then, the tilted operator could be computed from its definition 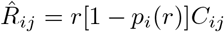. Candidate critical points were then detected as local minima of the smallest absolute eigenvalue of **L**(*r*), over a grid spanning Re *r* and Im *r*. Local minima on the grid were used as initial candidates and were refined by bounded local optimization using interpolation of the scanned response surface. Candidate singularities were retained only when the refined value was smaller than (10^*−*6^) and when the corresponding complex-conjugate candidate was also recovered within numerical tolerance. For each retained candidate, the activation mode was obtained from the near-null right eigenvector of **L**(*r*). Mode localization was quantified from the magnitude of the eigenvector components as explained in the main text.

### A.6 Respiratory pathogen in Italy

We constructed the province-level reproduction operator for Italy using Colocation Maps provided by Meta AI for Good [46], at the geographic resolution of provinces (ADM2, NUTS 3), for week 13 of 2023. In our analysis we also considered Italian regions, which are groupings of provinces and correspond to ADM1, NUTS 2 levels. Colocation Maps provide the probability 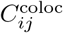 that a randomly chosen resident of province *i* and a randomly chosen resident of province *j* are located in the same 600 m × 600 m tile during a randomly chosen five-minute time window. Following the colocation-to-contact conversion used in previous work [33], we then parametrized **C** in Eq. (1) as 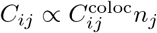, where *n*_*j*_ is the resident population of *j*, and normalized it to having spectral radius equal to one.

Importation risk was derived from estimated international air travel flows [47], following standard flow-based importation models [48, 49]. Specifically, let *W*_*ab*_ be the number of daily travelers from airport *a* to airport *b*. Let S be the set of airports in the source country and D_*i*_ be the set of airports in the province *i*. Then the importation risk to province *i* is 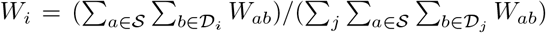. This implicitly assumes that prevalence is uniform in the source country, otherwise it can be easily adjusted to match nonuniform prevalence data [49].

Let us now consider vaccination. We assume that vaccines confer perfect and instantaneous immunity from infection. We do not consider roll-out delays nor behavioral changes following vaccination. We define a vaccination campaign as 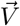, where *V*_*i*_ is the number of vaccinated individuals in province *i* (0 ≤ *V*_*i*_ ≤ *n*_*i*_), and *V*_*tot*_ = Σ_*i*_ *V*_*i*_ is the total stock of administered doses. Before the vaccination campaign, and for a given reproduction ratio, 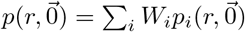 is the probability that one importation to the country leads to a large scale epidemic, where 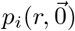 is the epidemic probability given introduction in province *i* (computed from Eq. (1) with *R*_*ij*_) and 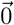 formally indicates a vaccination campaign with no doses administered. After the vaccination campaign, the proportion of susceptible residents in province *i* is then 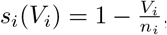, from which we can reconstruct the reproduction operator after the campaign as 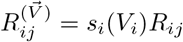, where *R*_*ij*_ is the reproduction operator before the campaign. We then recompute epidemic probabilities 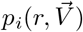. After the campaign, the probability that introduction to the country leads to an epidemic is 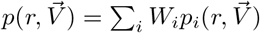. We then define the effectiveness of a vaccination campaign as the relative reduction in the probability that introduction leads to a large-scale epidemic, with respect to the scenario of no vaccination [50]: 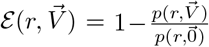. With this, we compared three vaccination strategies at fixed vaccine stock *V*_tot_ : uniform allocation, prioritization by importation risk, and prioritization informed by fragmented criticality. For the uniform strategy, vaccine doses were distributed proportionally to province population: 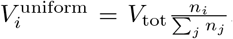. For the importation-based strategy, provinces were ranked by *W*_*i*_. For the fragmented-criticality strategy, we used the importation-risk score *W*_*i*_ as the baseline and reweighted it using the critical point information. For each province *i*, we associated it with the critical point for which its localization weight 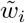 was maximal, and defined a *critical fragility index* as 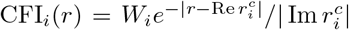. This score combines importation risk *W*_*i*_ with proximity to 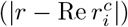 and strength of 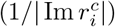 criticality.

For both targeted strategies, provinces were ranked by the corresponding score and the top *k* provinces were selected. In the CFI-based strategy, the province associated to the epidemic threshold (Im *r*^*c*^ = 0 so CFR → ∞) always ranked first. The fixed vaccine stock was then distributed among these selected provinces proportionally to their resident populations, with no province receiving more than one dose per resident. We varied *k* to obtain the curves shown in Fig. 2d.

### A.7 Measles in Texas

We constructed a county-level reproduction operator for Texas using Colocation Maps provided by Meta Data for Good [46], at the county level for week 9 of 2025, analogously to what was done for Italy. To incorporate susceptibility to measles, we used county-level school vaccination coverage from the Texas Department of State Health Services [36] and adjusted the reproduction operator analogously to what we did for the simulated vaccination experiment in Italy.

For each identified critical point (indexed *k*) we computed its normalized localization vector 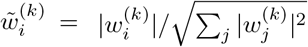 as previously described. Then, each county *i* was associated to the critical point that exhibited maximal localization on that county: 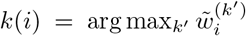. If multiple critical points gave equal weights within numerical tolerance, the county was assigned to that with the smaller imaginary part, i.e., the strongest. We compared the resulting activation-mode partition with three reference partitions: individual counties, seven broad administrative regions defined by the Texas Economic Development & Tourism Office, and metropolitan statistical areas based on the July 2023 delineations issued by the U.S. Office of Management and Budget [51, 52].

To quantify agreement between partitions, and specifically from the partition derived from fragmented criticality *C* and a reference partition *C*^*ref*^, we used B-cubed F1 score [53]. For a county *i*, let *C*_*i*_ be the set in *C* containing *i*, and the same for 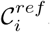. County level precision and recall are defined as 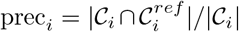 and 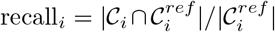. Average precision and recall are then simply Σ_*i*_ prec_*i*_ = prec */N* and recall = Σ_*i*_ recall_*i*_*/N*. Finally, B-cubed F1-score is customarily computed as *F* 1 = 2prec · recall*/*(prec + recall).

We used observed county-level measles burden during the 2025 Texas outbreak to test whether activation-mode groups are epidemiologically homogeneous [40]. Each county was assigned a binary burden class: *b*_*i*_ = 1 if county *i* reported at least one measles case, *b*_*i*_ = 0 otherwise. For a given partition *C*, we computed within-group burden agreement as a pair-counting homogeneity measure [54]: the fraction of within-group county pairs sharing the same burden class. For a group *c* ∈ *C*, this is 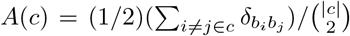. Groups with fewer than two counties were excluded from this pairwise calculation. The partition-level agreement was then computed as the pair-weighted average 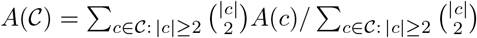. This statistic measures whether counties grouped together by a given partition tend to share the same outbreak status.

### A.8 Livestock-associated MRSA

We started from the One Health transmission model for livestock-associated methicillin-resistant S. aureus strain ST398 (LA-MRSA) published in Ref. [43]. The model contains eight epidemiological groups, making up the strata of the population: farmers (F), farm companion animals (FCA), transporters (T), slaughterhouse workers (SHW), veterinarians (V), companion animals (CA), the general human population (GH), and pig veterinarians (VP). Let *K*_*ij*_ be expected number of potentially infectious contacts generated in group *i* by one individual from group *j*, available in Ref. [43]. We then reconstructed the reproduction operator as usual as **R** = *r***K***/ρ*[**K**].

## Data availability

Meta Colocation Maps used for Italy and Texas can be requested through Meta Data for Good at https://dataforgood.facebook.com/dfg/tools/colocation-maps. Vaccination coverage, measles case counts, modeled air passenger flows, and LA-MRSA model parameters are available from the sources cited in the Methods. No new primary data were collected in this study.

## Code availability

The code used to compute the complex critical points and critical modes, perform the numerical analyses, and generate the figures in this study will be made publicly available at https://github.com/peasantxuan/Fragmented-criticality-of-infectious-disease-epidemics upon publication.

