## Supplementary material for "Fragmented criticality of infectious disease epidemics": Threshold_SI

### SUPPLEMENTARY MATERIAL
#### Fragmented criticality of infectious disease epidemics

Boxuan Wang[1] and Eugenio Valdano[1,*]

[1] *Sorbonne Université, INSERM, Sorbonne Public Health Institute,*
*Sorbonne Université Modeling Outbreaks Center, F75012, Paris, France.*

**2 Supplementary Figures** **7**

**Supplementary References** **10**

### 1 Supplementary Methods

#### 1.1 Generalizing to an arbitrary distribution of secondary infections

In the main text we assume a Poisson offspring distribution. Here, we relax this assumption and consider an arbitrary distribution

$$\Pr(X_{i\to j} = x) = f(x|R_{ji}), \tag{S1}$$

where $f(\cdot|\mu)$ is a one-parameter family of probability distributions with expectation $\mu$; additional parameters may be present but are assumed constant across strata. We denote its probability-generating function by

$$g(z|\mu) = \sum_{x=0}^{\infty} f(x|\mu) z^x. \tag{S2}$$

We prove here the steps necessary to show that the theory and findings of the main text, derived in the case of a Poisson distribution, hold in general.

Equation 1 of the main text generalizes to

$$F_i(\mathbf{p}, r) = p_i - 1 + \prod_j g\left(1 - p_j | r C_{ji}\right). \tag{S3}$$

**Reproduction operator and resolvent**

Equations 5 and 6 of the main text, before assuming a Poisson distribution, imply that the tilted probability mass and generating functions of the tilted process (conditioned on extinction) are

$$\hat{f}\left(x|R_{ij}\right) = \frac{f\left(x|R_{ij}\right)\left(1 - p_i\right)^x}{g\left(1 - p_i|R_{ij}\right)}; \tag{S4}$$

$$\hat{g}\left(z|R_{ij}\right) = \frac{g\left(z(1 - p_i)|R_{ij}\right)}{g\left(1 - p_i|R_{ij}\right)}. \tag{S5}$$

Consequently, the tilted reproduction operator is

$$\hat{R}_{ij} = \hat{g}'\left(1|R_{ij}\right) = \left(1 - p_i\right)\frac{g'\left(1 - p_i|R_{ij}\right)}{g\left(1 - p_i|R_{ij}\right)}. \tag{S6}$$

The derivation of $\mathbf{Y} = (1 - \hat{\mathbf{R}}^T)^{-1}\mathbf{1}$ in the main text already holds in general and does not depend on the Poisson assumption. The only requirement is to use the tilted reproduction operator from Eq. (S6).

**Relationship between critical points and bifurcations**

$$L_{ij} = \frac{\partial F_i}{\partial p_j} = \delta_{ij} - \frac{g'\left(1 - p_j|R_{ji}\right)}{g\left(1 - p_j|R_{ji}\right)}\prod_k g\left(1 - p_k|R_{ki}\right) \tag{S7}$$

$$= \delta_{ij} - \left(1 - p_i\right)\frac{g'\left(1 - p_j|R_{ji}\right)}{g\left(1 - p_j|R_{ji}\right)} = \delta_{ij} - U_{ij}, \tag{S8}$$

with

$$U_{ij} = \left(1 - p_i\right)\frac{g'\left(1 - p_j|R_{ji}\right)}{g\left(1 - p_j|R_{ji}\right)}. \tag{S9}$$

Comparing Eq. (S6) and Eq. (S9), one sees that the relationship between $\hat{\mathbf{R}}$ and $\mathbf{U}$ proven in the main text holds in general. Moreover, the form of the Jacobian $\mathbf{L}$ (Eq. (S8)) is also the same.

Also, the linearized evolution of $p(t)$ close to the fixed point is, assuming $\mathbf{p}(t) = \mathbf{p} + \mathbf{a}(t)$ as in the main text, exactly as in the main text:

$$\mathbf{a}(t+1) = \mathbf{U}\mathbf{a}(t). \tag{S10}$$

This completes the proof that the relationship between the complex-valued critical points and the bifurcation points of Eq. (S3) is the same as the one proven in the main text.

#### 1.2 Disappearance of fragmentation in homogeneous and strongly coupled populations

##### 1.2.1 Homogeneous transmission rates

Let us consider a matrix $\mathbf{C}$ defining the reproduction operator such that $\sum_j C_{ji} = 1$, i.e., column-stochastic. This means that the number of secondary infections that any individual generates (and whose expectation is $\sum_j R_{ji} = r \; \forall i$) is independent of the stratum of origin: Transmission potentials are homogeneous across strata [1]. In this case, we expect criticality not to be stratified and the epidemic threshold to describe the whole critical behavior of the system. We devote this section to proving that this is the case. Note that this condition is less restrictive than homogeneous mixing, because the repartition of secondary infections across strata can still depend on the stratum of origin. It does, however, include *homogeneous mixing* as the special case of $C_{ij} = 1/N$.

When $\mathbf{C}$ is column-stochastic, the solution to the equations defining epidemic probabilities $p_i = 1 - \exp(-r\sum_j C_{ji}p_j)$ is the same across strata: $p_i = p$ with $p = 1 - \exp(-rp)$. Therefore, $\mathbf{L} = 1 - \hat{\mathbf{R}}^T = 1 - r(1-p)\mathbf{C}^T$ and $\mathbf{U} = r(1-p)\mathbf{C}^T$. Let $\lambda_a$ denote the eigenvalues of $\mathbf{C}$ with $\lambda_1 = 1$ and let us assume that the inter-strata network is strongly connected so that $|\lambda_a| < 1$ for every non-Perron mode. A critical point $\tilde{r}$ associated with mode $\lambda_a$ other than the epidemic threshold (so $a \neq 1$) requires that $\tilde{r}[1 - p(\tilde{r})]\lambda_a = 1$. This means

$$\tilde{r}[1 - p(\tilde{r})] = \frac{1}{\lambda_a}. \tag{S11}$$

At the same time, the spectral radius $\rho$ of $\mathbf{U}$ is $\rho[\mathbf{U}] = |\tilde{r}[1 - p(\tilde{r})]|$.

$$\rho[\mathbf{U}] = |\tilde{r}\,[1 - p(\tilde{r})]| = \frac{1}{|\lambda_a|} > 1. \tag{S12}$$

This proves that $\tilde{r}$ is an unstable bifurcation point and thus not a critical point of the fragmented criticality.

Moreover, because $\mathbf{C}^T\mathbf{1} = \mathbf{1}$,

$$\bar{\mathbf{Y}} = \left[1 - r(1-p)\mathbf{C}^T\right]^{-1}\mathbf{1} = \frac{\mathbf{1}}{1 - r(1-p)}, \tag{S13}$$

so the expected size of a minor outbreak is independent of the stratum in which it is seeded. Homogeneity eliminates fragmented criticality: all non-collective singularities of the resolvent are unstable, and the admissible critical behavior is restricted to the single collective Perron mode.

##### 1.2.2 Strongly coupled strata

If strata are strongly coupled together, i.e., individuals from any stratum generate infections across all strata, we should expect the epidemic threshold to describe well the critical profile of the system. In other words, we expect fragmentation of criticality to progressively disappear as inter-stratum coupling increases.

It is not possible to find a general explicit solution for the strongly coupled regime as we did for the weakly coupled regime. It is possible, however, to observe it in the two-stratum model of Fig. 1 (main text). The trajectory of the critical points of that model as coupling increases illustrates the general pathway from fragmented criticality to a single critical point in strongly coupled populations.

We parametrize the model of Fig. 1 (main text) as $C_{11} = 1/m$ and $C_{21} = \gamma$. The resulting operator is

$$\mathbf{C} = \begin{pmatrix} 1/m & 0 \\ \gamma & 1 \end{pmatrix}. \tag{S14}$$

We consider $m > 1$, so that the epidemic threshold of the system is equal to one for any value of $\gamma$. We consider $\gamma \in [0, 1 - 1/m]$. At $\gamma = 0$ the system is decoupled, with $r_1^c = m$ (localized in 1) and $r_2^c = 1$ (localized in 2). For $\gamma = 1 - 1/m$, $\mathbf{C}$ is column-stochastic and the system is homogeneous (see Sec. 1.2.1).

At generic $\gamma$, the following expressions hold:

$$F_1 = p_1 - 1 + \exp\left[-r\left(\frac{p_1}{m} + \gamma p_2\right)\right] = 0; \tag{S15}$$

$$F_2 = p_2 - 1 + \exp(-r p_2) = 0; \tag{S16}$$

$$L_{11} = 1 - \frac{r}{m}\left(1 - p_1\right); \tag{S17}$$

$$L_{22} = 1 - r\left(1 - p_2\right). \tag{S18}$$

Equation (S16) is solvable at all $\gamma$, giving $p_2 = 1 + w/r$, with $w = \mathcal{W}_0(-re^{-r})$. Note that $\mathbf{L}$ is triangular so $L_{11}, L_{22}$ completely determine its spectrum. $L_{22} = 0$ identifies the bifurcation associated to the epidemic threshold $r_2^c$. We are interested in tracking the other critical point, given by $L_{11} = 0$, which gives $p_1 = 1 - m/r$. Inserting all this into Eq. (S15), one gets

$$\log\left(\frac{r}{m}\right) - \frac{r}{m} + 1 - \gamma(r + w) = 0. \tag{S19}$$

We know that for a range $\gamma \in [0, \gamma_\star]$, with $\gamma_\star \leq 1 - 1/m$ criticality is fragmented. At $\gamma = 1 - 1/m$ it is not fragmented because the system is homogeneous (Sec. 1.2.1). We posit the transition from fragmented criticality to *homogeneous* criticality to occur above a value $\gamma_\star$ for the coupling. If $\gamma_\star = 1 - 1/m$ then criticality is fragmented all the way until homogeneous population. If $\gamma_\star < 1 - 1/m$ fragmentation disappears before the system reaches homogeneous population.

In this simplified model, fragmentation disappears when the bifurcation identifying $r_1^c$ becomes unstable. This occurs if $|r(1 - p_2)| \geq 1$ (see Eq. (S18)). We thus impose the condition $|r(1 - p_2)| = 1$ to find the limiting coupling value $\gamma_\star$. Given the expression of $p_2$, this implies $|w| = 1$. We can thus parametrize $w = e^{i\theta}$, with $\theta \in (0, \pi]$. The expression of $p_2$ also implies $we^w = -re^{-r}$, which is equivalent to

$$\log r = r + w + i(\theta - \pi), \tag{S20}$$

where the branch of the logarithm is chosen so that $r = 1$ for $\theta = \pi$. All this can be used to derive an expression for $\gamma$ from Eq. (S19):

$$\gamma = \frac{\log(r/m) - r/m + 1}{r + w} = \frac{m - 1}{m} + \frac{1 - \log m + w/m + i(\theta - \pi)}{r + w}. \tag{S21}$$

The last condition to find $\gamma_\star$ is to find the value of $\theta = \theta_\star$ for which $\operatorname{Im}\gamma = 0$: in Eq. (S21). Specifically, we use the parametrization $r = \rho e^{i\phi}$. We need to find the values $\theta_\star, \rho_\star, \phi_\star$ that, in Eq. (S21), give $\operatorname{Im}\gamma = 0$

and thus $\gamma_\star$. They are the following. The first two are the real and imaginary parts of Eq. (S20), the last is $\operatorname{Im}\gamma = 0$:

$$\begin{cases} \log\rho_\star - \rho_\star\cos\phi_\star = \cos\theta_\star \\ \phi_\star - \rho_\star\sin\phi_\star = \theta_\star - \pi + \sin\theta_\star \\ \left(\theta_\star - \pi + \frac{\sin\theta_\star}{m}\right)\log\rho_\star = \left(1 - \log m + \frac{\cos\theta_\star}{m}\right)(\phi_\star - \theta_\star + \pi) \end{cases} \tag{S22}$$

The solution of the above system, inserted in Eq. (S21), gives the value of the coupling $\gamma_\star$ above which fragmentation disappears. Numerical evaluations of $\gamma_\star$ show that fragmentation can disappear at coupling lower than that of homogeneous population. For example, choosing $m = 2$ as in Fig. 1 of the main text gives $\gamma_\star \approx 0.22$, a value of coupling well below $\gamma = 1 - 1/m = 0.5$.

#### 1.3 An exactly solvable mean field

There is a simple mean-field model that is exactly solvable and illustrates the phenomenology under study. Let us suppose that there are $N$ distinct strata, each with a local reproduction ratio $r_i$. Instead of tracking specific between-strata connections $R_{ij}$ we link each of them to a mean field with a very large local reproduction ratio ($R_{mf,mf} \to \infty$). Each stratum can generate infections in the mean field with $R_{mf,i} = g_i$. The very large local reproduction ratio of the mean field means that infections generated in the mean field will always lead to major outbreaks: $p_{mf} \to 1$.

The epidemic threshold of the whole system is clearly $r = 0$ because the mean field is always active. Each stratum has an additional associated critical point. Since each stratum is independent from the other and only connected to the mean field, we can treat each stratum individually and drop the index $i$: $r_i \to r$, $g_i \to g$, $p_i \to p$. Note that, like this, this model analogous to the two-stratum model of Fig. 1**a** of the main text, with a very large local transmissibility in the receiving stratum ($R_{22} \to \infty$ in Fig. 1a of the main text).

Under these conditions the equation of $p$

$$p = 1 - e^{-rp - gp_{mf}} = 1 - e^{-rp-g} \tag{S23}$$

has the following explicit solution:

$$p = 1 + \frac{1}{r}\mathcal{W}_k\left(-re^{-r-g}\right) \tag{S24}$$

$$\text{with } k = 0, -1 \;. \tag{S25}$$

$\mathcal{W}_0$ is the principal branch of the Lambert's W function and $\mathcal{W}_{-1}$ is the only other branch containing the real axis. The reproduction operator is lower triangular because each stratum is connected to the mean field only (and not viceversa) and the diagonal entry for the mean field is trivial, and the same holds for the tilted reproduction operators. So again each stratum can be treated individually, through the resolvent

$$\left(1 - \hat{\mathbf{R}}\right)^{-1} = [1 - (1-p)r]^{-1} = \left[1 + \mathcal{W}_k\left(-re^{-r-g}\right)\right]^{-1} . \tag{S26}$$

The resolvent is singular if $\mathcal{W}_k(-re^{-r-g}) = -1$, which requires

$$-r_c e^{-r_c - g} = -e^{-1}, \tag{S27}$$

which corresponds to the branch point of the Lambert's W function. The singularity of the resolvent is located where the two solutions of Eq. (S23), corresponding to the $0, -1$ branches of W, overlap, i.e., in the bifurcation point of the solutions, confirming the correspondence between critical points and bifurcation points in this mean field model too, here by means of the branch properties of the Lambert's W function.

Equation (S27) can also be solved explicitly by means of the Lambert's W function:

$$r_c = -\mathcal{W}_h(-e^{g-1}). \tag{S28}$$

Equation (S28) is the exact, explicit form of the critical points of this mean field model, valid for any value of coupling $g$. If the stratum is decoupled from the mean field ($g = 0$) the solution is, expectedly, unique and real: $r_c = 1$. It is the epidemic threshold of the isolated stratum. If $g > 0$, there exist two non-real, complex conjugate ($h = 0, -1$) solutions, which represent the smeared critical point related to the stratum under the effect of the mean field.

Assuming small coupling, one can make them even more explicit and link them to the weak coupling expansion of the main text:

$$r_c = -\mathcal{W}_h(-e^{g-1}) = 1 \pm i\sqrt{2g} - \frac{2g}{3} + \mathcal{O}\left(g^{3/2}\right). \tag{S29}$$

Figure S1 plots the critical point in Eq. (S29) as a function of the coupling $g$, displaying the two complex-conjugate points arising from the $h = 0, -1$ branches of W and the accurate performance of the weak coupling expansion up to linear order in $g$.

#### 1.4 Generalizing to degenerate critical eigenspaces

The derivations in the main text assume that the eigenvalue becoming critical is nondegenerate. Here, we show that the construction extends directly to a critical eigenspace of dimension larger than one.

##### Critical points

Let $(\mathbf{p}^c, r^c)$ be a critical point at which $\dim\ker\mathbf{L} = m$. Since $\mathbf{L}$ is square, its cokernel has the same dimension. Let $\{\mathbf{u}_a\}_{a=1}^m$ be a basis of $\ker\mathbf{L}$ and decompose

$$\mathbf{p} = \mathbf{p}^c + \sum_{a=1}^{m} \xi_a \mathbf{u}_a + \mathbf{q}, \tag{S30}$$

where $\mathbf{q}$ belongs to a complementary subspace. The Lyapunov–Schmidt reduction proceeds exactly as in the nondegenerate case. Projecting $\mathbf{F} = 0$ onto Im$\mathbf{L}$ determines $\mathbf{q} = \mathbf{q}(\boldsymbol{\xi}, r)$, because the restriction of $\mathbf{L}$ to the complementary subspace is invertible. Projecting the remaining equations onto the cokernel gives an $m$-dimensional reduced system

$$\mathbf{G}(\boldsymbol{\xi}, r) = 0, \tag{S31}$$

instead of the single scalar equation obtained for $m = 1$. In a basis adapted to the kernel and its complement, the same Schur-complement argument used in the main text gives

$$\det\mathbf{L} = \det\mathbf{L}_{\perp\perp} \det\left(\frac{\partial\mathbf{G}}{\partial\boldsymbol{\xi}}\right), \tag{S32}$$

with $\det\mathbf{L}_{\perp\perp} \neq 0$. Thus, the condition defining the critical points is simply generalized from the vanishing derivative of a scalar reduced equation to the singularity of the $m$-dimensional reduced Jacobian. No further modification of the Lyapunov–Schmidt construction is required.

**Localization**

Let the critical eigenvalue $\lambda_c$ of $\hat{\mathbf{R}}$ have algebraic and geometric multiplicities equal to $m$, with $\lambda_c = 1$ at the critical point, and let $\{\mathbf{v}_a\}_{a=1}^m$ and $\{\mathbf{w}_a^T\}_{a=1}^m$ be bases of its right and left eigenspaces, respectively, normalized such that $\mathbf{w}_a^T \mathbf{v}_b = \delta_{ab}$. The singular contribution to the resolvent is

$$\left(1 - \hat{\mathbf{R}}^T\right)^{-1} \approx \frac{1}{1-\lambda_c} \sum_{a=1}^{m} \mathbf{w}_a \mathbf{v}_a^T. \tag{S33}$$

Consequently, the singular contribution to the expected outbreak size is

$$\mathbf{Y} \approx \frac{1}{1-\lambda_c} \sum_{a=1}^{m} h_a \mathbf{w}_a, \qquad h_a = \sum_i v_{a,i}. \tag{S34}$$

Although the individual eigenvectors are not uniquely defined within a degenerate eigenspace, their combined contribution in Eq. (S34) is. To show this explicitly, consider a change of basis

$$\mathbf{v}'_a = \sum_c A_{ac} \mathbf{v}_c, \qquad \mathbf{w}'_a = \sum_c B_{ac} \mathbf{w}_c. \tag{S35}$$

Preserving the normalization requires

$$\mathbf{w}'^{\,T}_a \mathbf{v}'_b = \sum_{c,d} B_{ac} A_{bd} \mathbf{w}_c^T \mathbf{v}_d = \sum_c B_{ac} A_{bc} = \delta_{ab}, \tag{S36}$$

and therefore $\mathbf{B} = \mathbf{A}^{T,-1}$. The operator entering Eq. (S33) is then invariant:

$$\sum_a \mathbf{w}'_a \mathbf{v}'^{\,T}_a = \sum_{a,c,d} B_{ac} A_{ad} \mathbf{w}_c \mathbf{v}_d^T \tag{S37}$$

$$= \sum_{c,d} \left(\mathbf{B}^T \mathbf{A}\right)_{cd} \mathbf{w}_c \mathbf{v}_d^T \tag{S38}$$

$$= \sum_c \mathbf{w}_c \mathbf{v}_c^T. \tag{S39}$$

Thus, the intrinsic object associated with a degenerate critical point is not any individual eigenvector but the spectral projector onto the entire critical eigenspace,

$$\mathbf{\Pi}_c = \sum_a \mathbf{w}_a \mathbf{v}_a^T. \tag{S40}$$

Its action on the source vector $\mathbf{1}$ determines the effect of the critical point on the expected outbreak size:

$$[\mathbf{\Pi}_c \mathbf{1}]_i = \sum_a h_a w_{a,i}. \tag{S41}$$

This quantity is basis independent and therefore provides the natural generalization of the localization measure used in the main text. The relative sensitivity of stratum $i$ to a degenerate critical point can be defined as

$$\frac{\left|[\mathbf{\Pi}_c \mathbf{1}]_i\right|}{\sqrt{\sum_j \left|[\mathbf{\Pi}_c \mathbf{1}]_j\right|^2}}. \tag{S42}$$

For $m = 1$, this expression reduces to $|w_i|/\sqrt{\sum_j |w_j|^2}$, up to the overall normalization of the eigenvectors.

### 2 Supplementary Figures

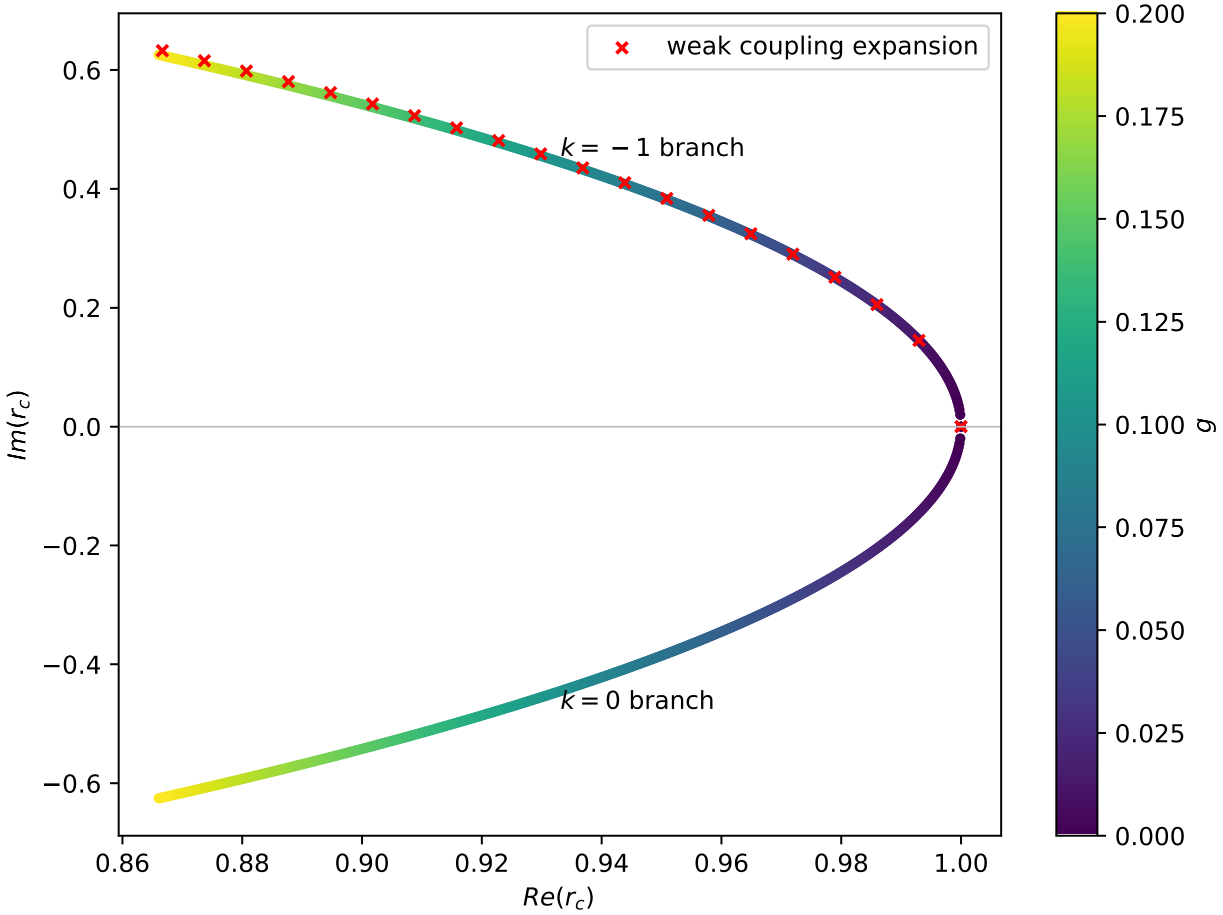

**Fig.S1**|Critical point of the mean field model. The axes display the real and imaginary parts of the critical point of the mean field model in Eq. (S29). The two complex conjugate points arising from the two different branches of the Lambert's W are visible, as well as the weak coupling expansion up to linear order in $g$.

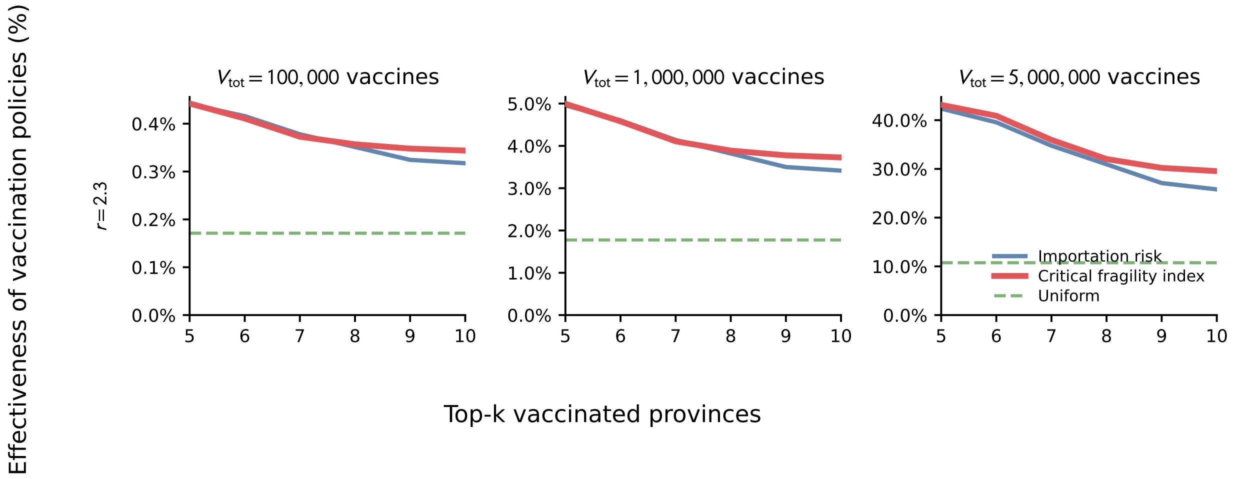

**Fig.S2**|Effectiveness of vaccination policies under an importation scenario from China. Effectiveness of vaccination policies in the simulated experiment of preventive vaccination: uniform allocation (green dashed), allocation based on risk of importation (blue), and allocation based on critical fragility index (red). Columns correspond to vaccine stocks of $V_{\text{tot}} = 10^5$, $10^6$, and $5 \times 10^6$ doses. The assumed reference reproduction ratio is $r = 2.3$, consistent with estimates of $R_0 = 2.33$ during the early SARS-CoV-2 epidemic in China [2]. The horizontal axis gives the number of provinces prioritized for vaccination. The vertical axis reports the estimated effectiveness, as defined in the Methods section "Respiratory pathogen in Italy" of the main paper.

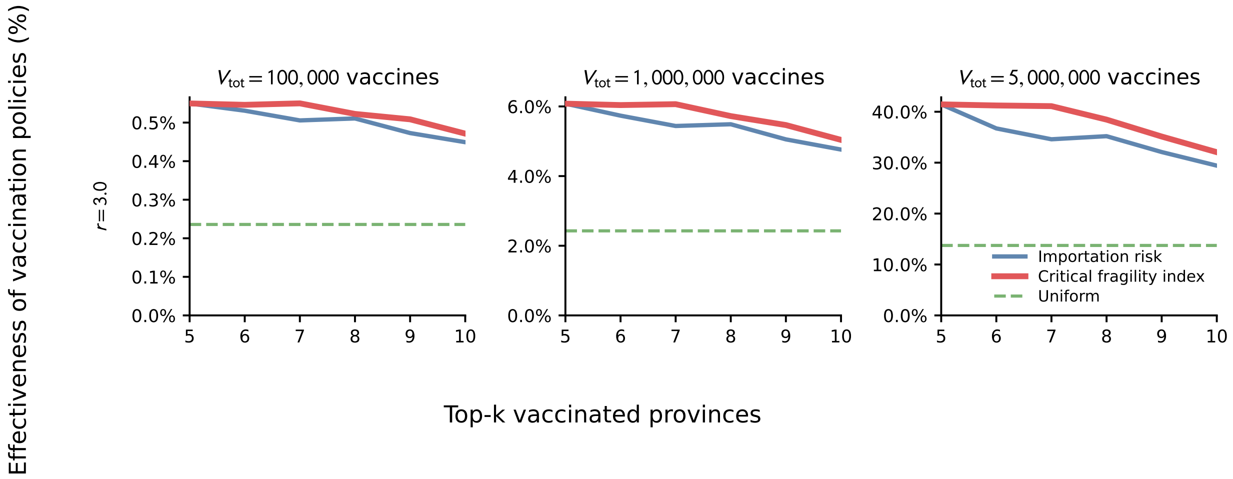

**Fig.S3**|Effectiveness of vaccination policies under an importation scenario from Botswana. Effectiveness of vaccination policies in the simulated experiment of preventive vaccination: uniform allocation (green dashed), allocation based on risk of importation (blue), and allocation based on critical fragility index (red). Columns correspond to vaccine stocks of $V_{\text{tot}} = 10^5$, $10^6$, and $5 \times 10^6$ doses. The assumed reference reproduction ratio is $r = 3.0$, corresponding approximately to a 30% increase in transmissibility relative to the early SARS-CoV-2 reference value $R_0 = 2.33$ [2, 3]. Botswana was considered as a representative southern African importation scenario, motivated by the rapid epidemic expansion of the SARS-CoV-2 Omicron variant in southern Africa [4]. The horizontal axis gives the number of provinces prioritized for vaccination. The vertical axis reports the estimated effectiveness, as defined in the Methods section "Respiratory pathogen in Italy" of the main paper.
